# Demographic, spatial, and temporal dietary intake patterns among 526,774 23andMe research participants

**DOI:** 10.1101/2020.04.14.20058263

**Authors:** Janie F. Shelton, Briana Cameron, Stella Aslibekyan, 23andMe Research Team, Robert Gentleman

## Abstract

**Objective:** To characterize dietary habits, their temporal and spatial patterns, and associations with body mass index (BMI) in the 23andMe study population.

**Design:** We present a large-scale cross-sectional analysis of self-reported dietary intake data derived from the web-based NHANES 2009-2010 dietary screener. Survey-weighted estimates for each food item were characterized by age, sex, race/ethnicity, education, and BMI. Temporal patterns were plotted over a 2-year time period, and average consumption for select food items was mapped by state. Finally, dietary intake variables were tested for association with BMI.

**Setting:** U.S. based adults 20-85 years of age participating in the 23andMe research program.

**Participants:** Participants were 23andMe customers who consented to participate in research (n=526,774) and completed web-based surveys on demographic and dietary habits.

**Results:** Survey-weighted estimates show very few participants met federal recommendations for fruit: 2.6%, vegetables: 5.9%, and dairy intake: 2.8%. Between 2017-2019, fruit, vegetables, and milk intake frequency declined, while total dairy remained stable and added sugars increased. Seasonal patterns in reporting were most pronounced for ice cream, chocolate, fruits, and vegetables. Dietary habits varied across the U.S., with higher intake of sugar and calorie dense foods characterizing areas with higher average BMI. In multivariate-adjusted models, BMI was directly associated with intake of processed meat, red meat, dairy, and inversely associated with consumption of fruit, vegetables, and whole grains.

**Conclusions:** 23andMe research participants have created an opportunity for rapid, large scale, real time nutritional data collection, informing demographic, seasonal and spatial patterns with broad geographical coverage across the U.S.

## Introduction

Inadequate intake of healthy foods (fruits, vegetables, whole grains, nuts/seeds) and excessive intake of foods high in sodium, added sugars, and saturated fat are major contributors to excess morbidity and premature mortality worldwide.^(1, 2)^ In the United States, approximately half of all cardio-metabolic deaths are attributable to suboptimal intake of fruits, vegetables, nuts, seeds, and whole grains and excess consumption of salt, processed meats, and sugary beverages^(3)^ and per the dietary guidelines outlined by the American Heart Association to maximize cardiovascular health, 45.6% of Americans adults are estimated to have a “poor diet”, with less than 10% consuming adequate amounts of fruits and vegetables.^(4)^

Poor dietary habits and inadequate physical activity are major drivers of elevated body mass index (BMI), which in turn increases the risk of developing adverse cardio-metabolic outcomes.^(5)^ Diet and physical activity also represent the most actionable areas at both the individual and population levels to prevent chronic disease.^(6)^ While the majority of dietary recommendations are geared towards average effects with measurable benefits to population health, the science of precision nutrition has been uncovering informative subgroup variation.

For example, among those who consume 22g/day or more of saturated fat, weight gain was more pronounced among those with the -265 C/C genotype of *APOA2*, an estimated 10-20% of the population, compared to those without it.^(7)^ However, longer term health consequences, such as varying cardio-metabolic disease risk across combinations of exposures, are not well understood. While precision nutrition has been described as a major priority for epidemiology,^(8)^ the vast majority of studies published to date are underpowered for more granular discovery.

In order to advance our understanding of how nutritional and other factors interact and impact health, data are needed from large populations followed over time. Since 2017, over 500,000 genotyped 23andMe research participants answered a survey about their dietary habits over the past month. Due to the growing size of the customer database, widespread geographical representation, and continuous data collection over time, data provided by 23andMe research participants now represent a large enough sample to inform population-based inferences for a variety of health behaviors, including diet.

However, because 23andMe participants are a subsample of 23andMe personal genome service customers, they are not a representative sample of the general population. Therefore, the aim of this manuscript is to describe how the sociodemographic profile and dietary habits of the 23andMe research participants compare to the US population, to characterize its dietary habits using survey weights that account for potential imbalances, and to test cross-sectional associations between BMI and consumption of several food items in this uniquely large cohort. Our manuscript illustrates both the potential and the caveats of conducting nutritional epidemiology research in large-scale digital cohorts.

## Methods

This study used data from consented 23andMe research participants from the U.S. aged 20 years or older who completed the NHANES 2009-2010 dietary screener questionnaire on the 23andMe website or mobile application.

This study was conducted according to the guidelines laid down in the Declaration of Helsinki and all procedures involving research study participants were approved by the Ethical & Independent Review Services, a private independent institutional review board. All 23andMe customers consent to participate in research online and consent is captured electronically. Ethical & Independent Review Services approved this form of informed consent and waived the requirement to obtain signed consent under U.S. law 45 CFR 46.117(c).

Customer data are de-identified; 23andMe researchers who conducted the statistical analyses in this manuscript did not have access to personally identifiable information (e.g. name, address, etc) and were trained in the responsible conduct of research. 23andMe has obtained a Certificate of Confidentiality from the National Institutes of Health, further protecting the privacy of research participants. Additional consent form information is available at www.23andme.com/about/consent.

### Data collection

Once a 23andMe personal genome service customer receives their sample collection kit, they are asked to register it online prior to return. During this process, all customers are invited to participate in research, which occurs predominantly through web-based research surveys, some of which are developed in-house by 23andMe and others are implemented using previously validated instruments. Beginning in March 2017, dietary intake frequencies were collected using a validated web-based version of the self-administered 25-item NHANES dietary screener questionnaire (2009/2010) otherwise referred to as the DSQ.^(9)^ Accompanying surveys provided self-reported data on covariates such as age, sex, race/ethnicity, education, and BMI. The questions on race/ethnicity and education utilized the same response options as the U.S. Census. Participant recruitment to the surveys took place through two approaches, actively through email and passively through the website and mobile application. Passive recruitment on the website occurs via the ‘research stream’, which is a feature on the 23andMe website and within the 23andMe app (available on both iOS and Android devices) which continuously surfaces surveys to eligible respondents on a variety of topics.

As such, the DSQ was fielded to those who had likely already completed higher priority surveys such as the Health survey (which collects data on basic demographics, health and disease status), or disease specific surveys based on conditions they may have reported. Because of this passive recruitment targeting scheme, we do not know the total population of people who were offered the survey, and therefore cannot directly estimate a survey response rate.

### Dietary assessment and intake estimation

The DSQ measures intake of fruits and vegetables (cups/day), dairy (cups/ day), calcium (mg), added sugars (g), whole grains (ounce equivalents), and fiber (g).^(10)^ Total fruit includes both whole fruit and fruit juice, vegetable intake is estimated from consumption frequency of salad, potatoes, beans, tomato sauce, salsa, pizza toppings, and other vegetables. Dairy intake is estimated from milk, cheese, ice cream, and pizza. Whole grains are derived from cereals, whole grain bread, popcorn, and whole grain rice. Added sugars are derived from soda, fruit drinks, cookies/cakes/pie, doughnuts, ice cream, added sugar/honey in coffee or tea, candy, and cereal. Added sugars from sugar sweetened beverages are derived from soda, fruit drinks, and sugar/honey added to coffee or tea.^(10)^

The DSQ and scoring algorithms to derive broader food groups were validated against dietary intake using the 24-hour recall method in a representative population of non-institutionalized U.S. based NHANES study participants aged 2-69.^(11)^ In that context, the DSQ was reported to produce stable estimates of intake for this set of dietary factors and correlate well with 24-hour recall estimates in three external study populations.^(11)^ In the DSQ, individual food items are ascertained as frequency over the past month. Based on the data processing and scoring procedures recommended for use with the DSQ, we converted monthly estimates to daily estimates, and then multiplied the latter by item-specific portion and serving size estimates provided according to age and sex.^(12)^ Components of cereals such as sugar, fiber, and whole grain content were derived from the classifications on a per cereal basis provided by NHANES.

To evaluate the percent of the population meeting dietary recommendations, we estimated the proportion of participants within strata of age and sex who met the United States Department of Agriculture’s Dietary Guidelines for Americans 2015-2020 recommended intake of fruits and vegetables.^(13)^

### Time trends and maps

Mean daily intake and 95% confidence intervals were plotted by week of survey completion to explore seasonal variation in dietary intake. Although the data collected refer to the past month, we did not apply a lag period or adjust the date in any way. Broader temporal changes in reported dietary habits over time are visualized with loess curves.

Participants self-reported their current zip code, which we subsequently mapped to states to characterize the geographic distribution of average intake of food groups, select food items, and average BMI across the U.S. Regional average intake was estimated for the South (Delaware, District of Columbia, Florida, Georgia, Maryland, North Carolina, South Carolina, Virginia, West Virginia, Alabama, Kentucky, Mississippi, Tennessee, Arkansas, Louisiana, Oklahoma, Texas), West (Arizona, Colorado, Idaho, New Mexico, Montana, Utah, Nevada, Wyoming, Alaska, California, Hawaii, Washington, and Oregon), Northeast (Connecticut, Maine, Massachusetts, New Hampshire, Rhode Island, Vermont, New Jersey, New York, Pennsylvania), and the Midwest (Indiana, Illinois, Michigan, Ohio, Wisconsin, Iowa, Kansas, Minnesota, Missouri, Nebraska, North Dakota, and South Dakota).

For this analysis, the foods selected for seasonality were those with the most pronounced seasonal trends: chocolate, fruit, ice cream and salad. Those selected for mapping showed the most pronounced geographic patterns: fruit, vegetables, whole grains, red meat, processed meat, dairy, and pizza.

### Development of survey weights

To address differences between our respondent population and the national population (Table 1), we developed survey weights and applied them to our sample. We used iterative proportional fitting (IPF) to calculate weights based on age (20-64, 65+), sex (M, F), race (white, non-white), education (less than college, completed college), and BMI (obese, not obese). We weighted our sample proportion to match the demographic distribution as reported by the U.S. Census (age, sex, race, education)^(14)^ and the Centers for Disease Control and Prevention.^(15)^ We excluded respondents (n=156, 053) from the raking procedure (implemented via the survey package (version 3.29.5)^(16)^ in R) and subsequent analyses if they did not provide this demographic information. To address the increase in standard error that may be introduced with large sample weights, we chose binary classifications instead of utilizing all strata (e.g. for BMI), and trimmed all initial weights to be less than 5-times the mean survey weight.^(17)^

**Table 1:**
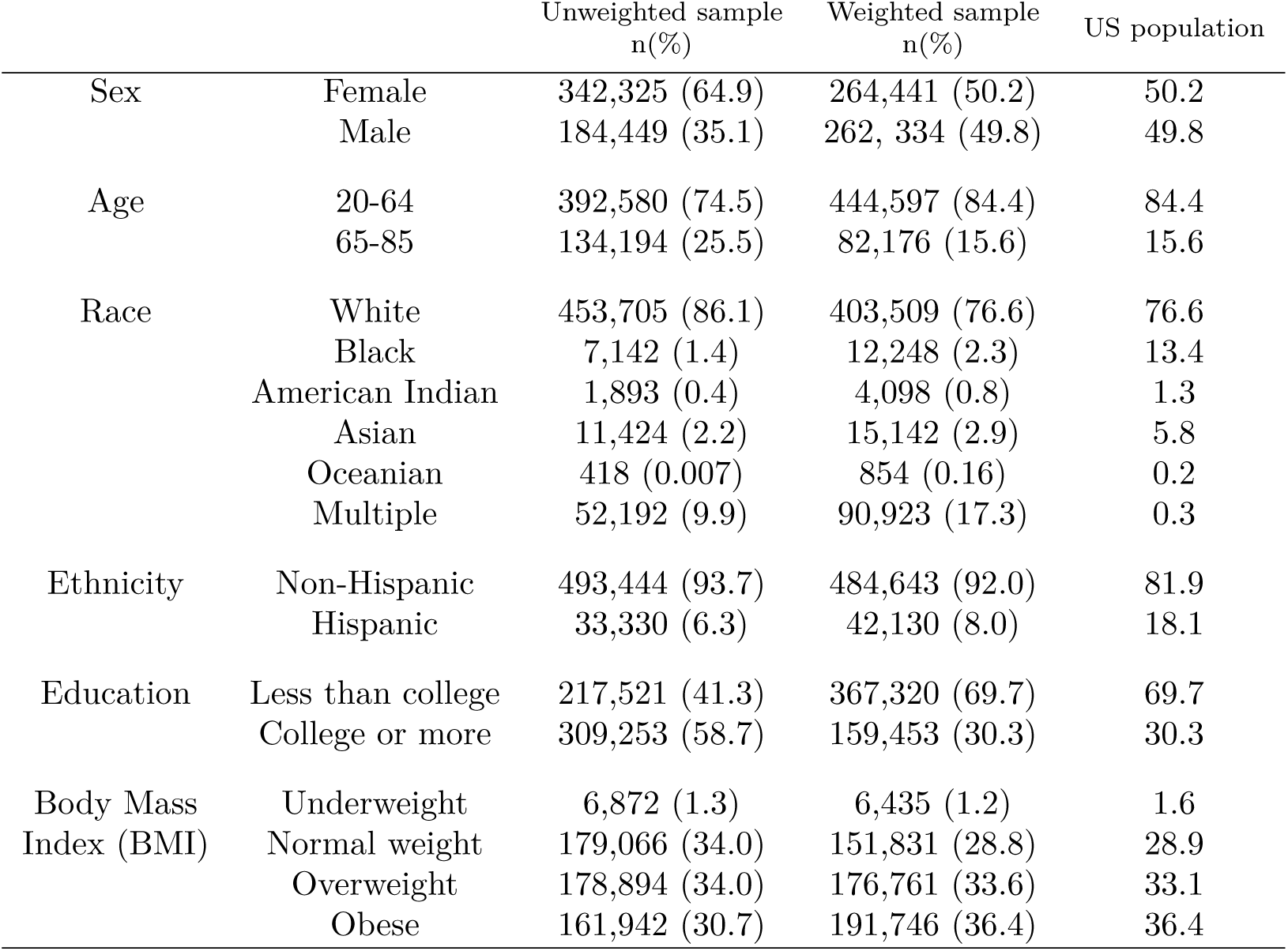
Respondent characteristics with complete data on age, sex, education, race/ethnicity, and body mass index compared to the national population drawn from the US Census.

### Dietary intake associations with BMI

To measure the cross-sectional correlation between dietary intake and BMI, we explored the association between each food item continuously using the daily frequency or estimated quantity for derived measures, but ultimately classified intake by tertiles for ease of interpretation and standardization of quantity (high vs. low intake frequency). We limited our sample to participants who self-reported height and weight values yielding estimated BMI scores between 14 and 70. We evaluated BMI both with and without log transformation.

During model development, race/ethnicity, education, age, sex, age*sex, and age squared were evaluated as potential confounders. The final linear models tested the associations between food intake frequency (highest vs. lowest consumption tertiles for all food items measured in the DSQ) and log-transformed BMI, adjusting for race/ethnicity, education, age (centered at 50 years), sex, and centered age squared to maximize the variance explained by the model and the uniformity of the plotted residuals. We estimated regression coefficient estimates and 95% confidence intervals for each food item to evaluate the relationship between frequency of intake and BMI.

We used R (R Foundation for Statistical Computing, version 3.2.5) for all statistical analyses and data visualization.

## Results

### Respondent characteristics

A total of 526,774 U.S.-based respondents with non-missing data for sex, age, education, race/ethnicity, and BMI completed the DSQ between March 2017 and August 2019. Compared to the U.S. population, sample respondents were more likely to be female (65% vs 50%), more likely to be white (86% vs. 77%), less likely to be Hispanic (6% vs. 18%), nearly twice as likely to have completed college (59% vs. 30%), and less likely to be obese (31% vs. 36%).

Application of sampling weights yielded a more representative sample based on age, sex, education, and BMI.

### Temporal characteristics of dietary intake

Due to the high-rate of gift giving of the 23andMe genetic testing kit during the holidays, survey completion between late December and early January is on average 4-5 fold greater than the average of other weeks throughout the year. Due to this high degree of response in these months, the precision of the estimates by season are highest over the Northern Hemisphere winter period as compared to other seasons (Figure 1).

**Figure 1:**
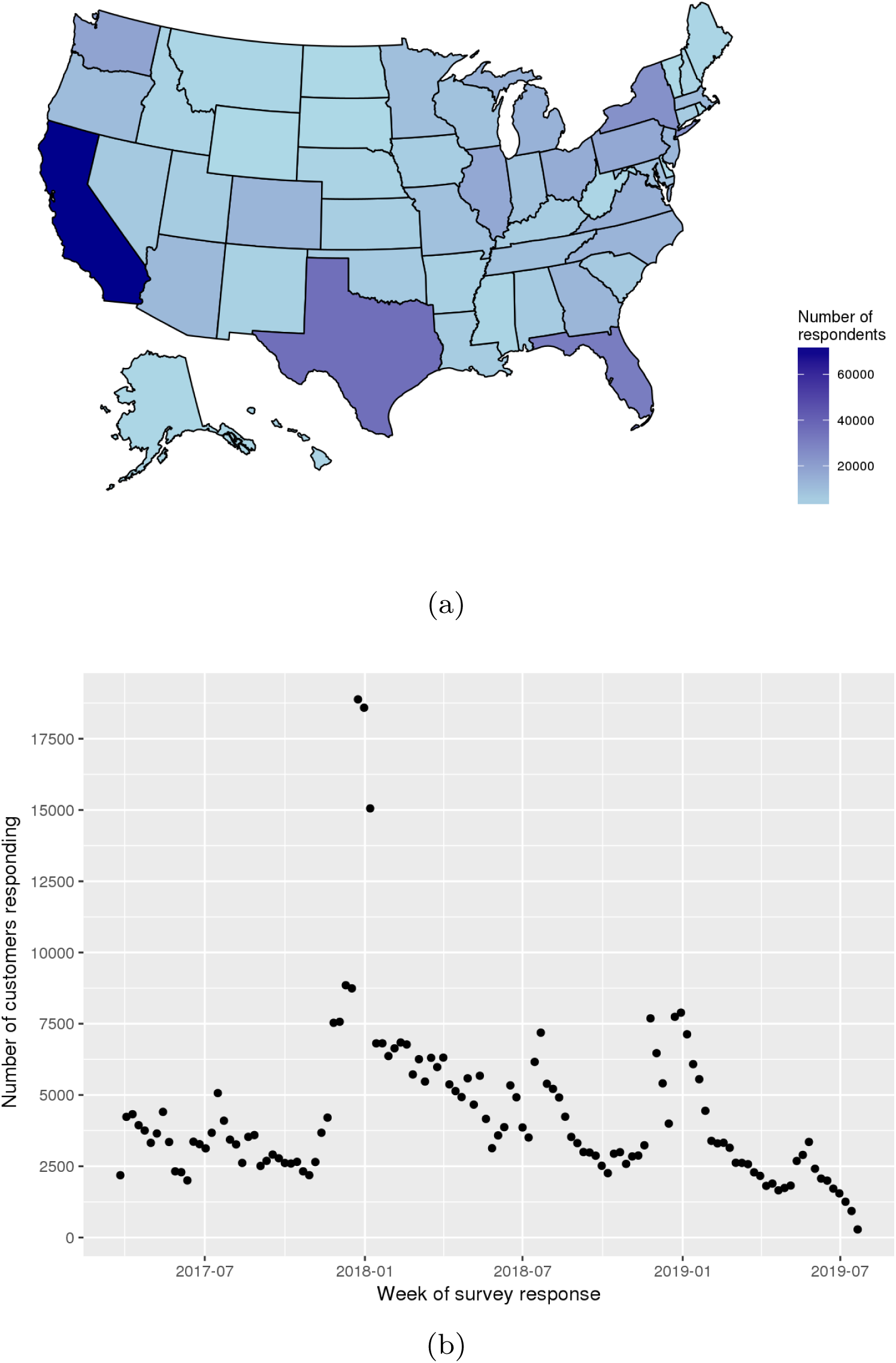
Survey completions by state (a) and week (b) during interval of data collection.

Because survey data have been collected continuously over two years, seasonal trends in dietary frequencies are observable. Figure 2 shows the unadjusted mean daily intake patterns for fruit, vegetables, salad, chocolate, and ice cream. Chocolate and ice cream show clear seasonal patterns, with peak chocolate consumption (0.42 times per day) in December-January compared to the lowest consumption in mid-June of 0.31 times per day. Peak ice cream consumption is observed in June-August, with an average frequency of 0.17 times per day compared to 0.01 times per day in February.

**Figure 2:**
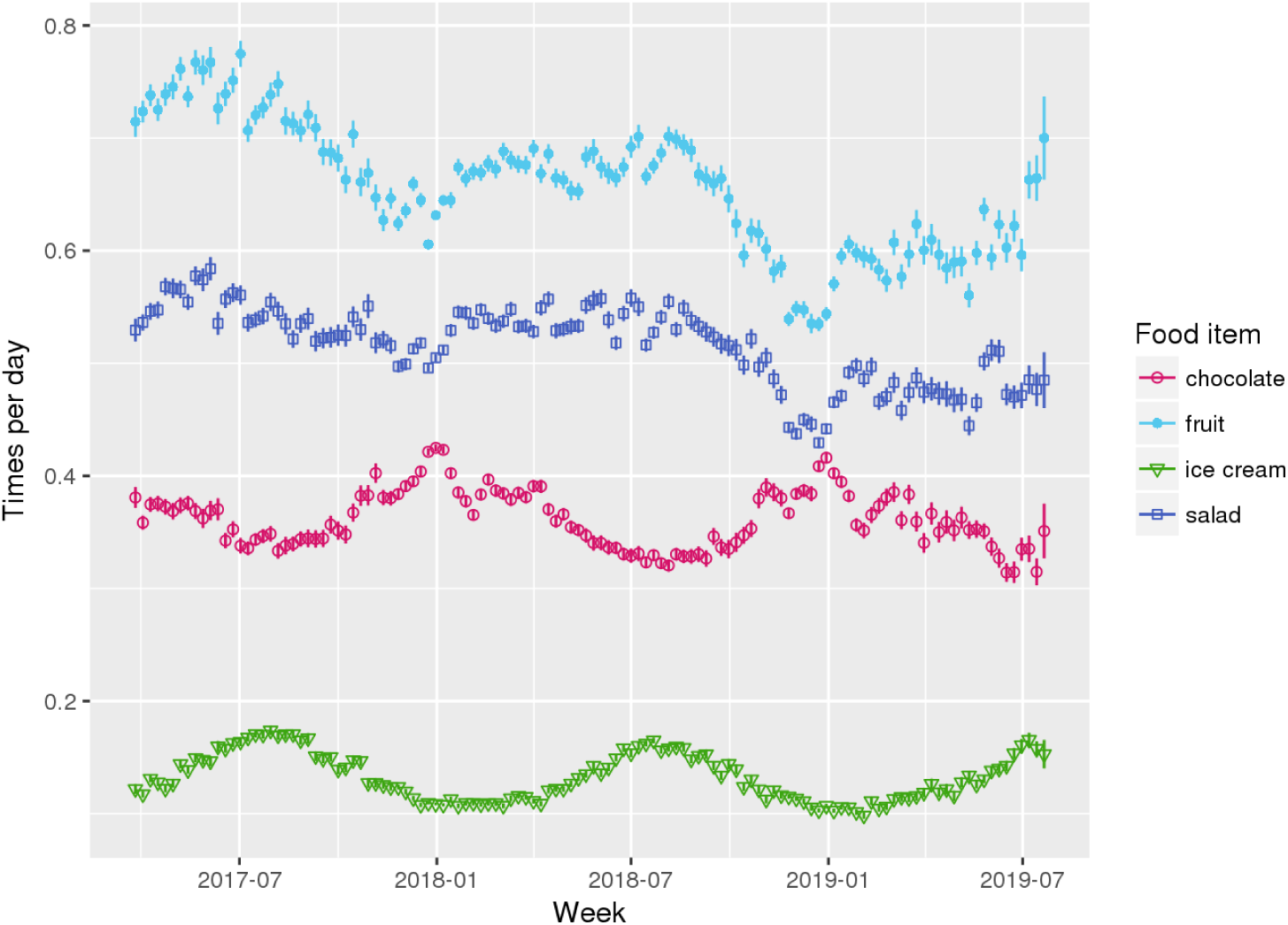
Seasonality patterns of chocolate, fruit, ice cream, and salad average daily intake by week of survey completion, 2017-2019.

Milk consumption, which includes both soy and dairy, declined by approximately 28% between 2017 and 2019 following a pattern evident since the 1970’s ^(17)^ and observed in the NHANES study between 1992-2001.^(19)^ However, only a very minimal decline in total dairy consumption is observed over time, likely owing to observed increases in cheese and pizza consumption (Figure S1).

For fruits and vegetables, we observed higher reported consumption in Northern Hemisphere summer than winter (Figure 2), but also noted a general decline over the reporting period (Figure 3). We have explored various possible explanations for this observation, such as changing customer demographics (e.g. age, sex, or type of genotyping kit purchased or the influx of winter customers), but saw no clear explanatory patterns. Further, because national published estimates are not yet available for this time period, we have no comparison on which to support or refute the observation that fruit and vegetable consumption is declining in the general population.

**Figure 3:**
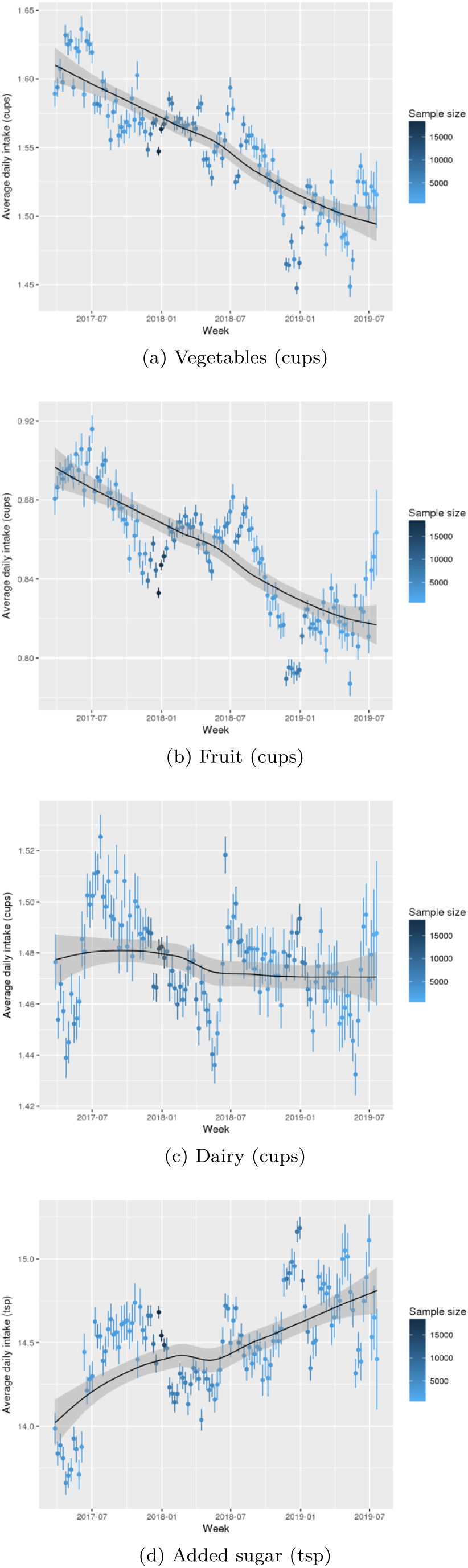
Mean and 95% confidence intervals with loess curves of the of intake frequency reported by week of data collection for vegetables, fruit, dairy, and added sugars, over the two-year data collection period (2017-2019).

### Spatial characteristics of dietary intake

The respondent population show high geographical coverage across the U.S. (Figure 1), with a minimum of 1,000 respondents in every state. Higher proportions of respondents were from high population density states (California, Texas, Florida). By exploring the fraction of respondents as a percent of the total adult population, we observed relative overrepresentation in California, Florida, and Pennsylvania and underrepresentation in Nebraska, South Dakota, and Vermont.

Minor regional differences are noted when comparing data in aggregate (Table 4), but intake frequencies plotted at the state level show distinct dietary patterns by region. In the Southeastern states, fruit, vegetable and whole grain intake are markedly lower than in coastal areas, and intake of processed meat, regular soda, and added sugars are comparatively higher. Higher average intake of milk, pizza, and red meat are observed in the northern Midwestern states, and while lower vegetable intake is observed, fruit and whole grain intake are similar to coastal areas (Figure 3). Dietary frequencies which correspond with higher BMI in Figure 4 are also demonstrated to spatially correspond with patterns of higher BMI at the state level (e.g. higher frequency of processed meat and lower consumption of fruits, vegetables, and whole grains in the southern U.S. where BMI is highest).

**Table 4:**
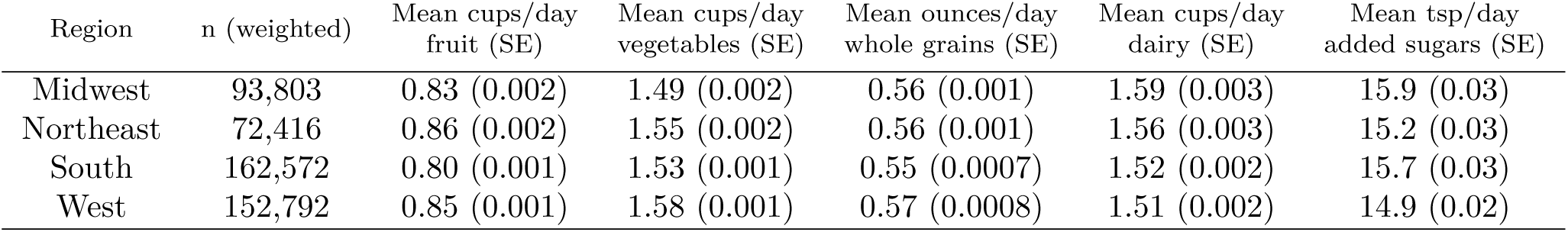
Survey-weighted mean (SE) intake of select vegetables, fruit, dairy, added sugars, and whole grains by region (2017-2019).

| Region | n (weighted) | Mean cups/day<br>fruit (SE) | Mean cups/day<br>vegetables (SE) | Mean ounces/day<br>whole grains (SE) | Mean cups/day<br>dairy (SE) | Mean tsp/day<br>added sugars (SE) |
| --- | --- | --- | --- | --- | --- | --- |
| Midwest | 93,803 | 0.83 (0.002) | 1.49 (0.002) | 0.56 (0.001) | 1.59 (0.003) | 15.9 (0.03) |
| Northeast | 72,416 | 0.86 (0.002) | 1.55 (0.002) | 0.56 (0.001) | 1.56 (0.003) | 15.2 (0.03) |
| South | 162,572 | 0.80 (0.001) | 1.53 (0.001) | 0.55 (0.0007) | 1.52 (0.002) | 15.7 (0.03) |
| West | 152,792 | 0.85 (0.001) | 1.58 (0.001) | 0.57 (0.0008) | 1.51 (0.002) | 14.9 (0.02) |

**Figure 4:**
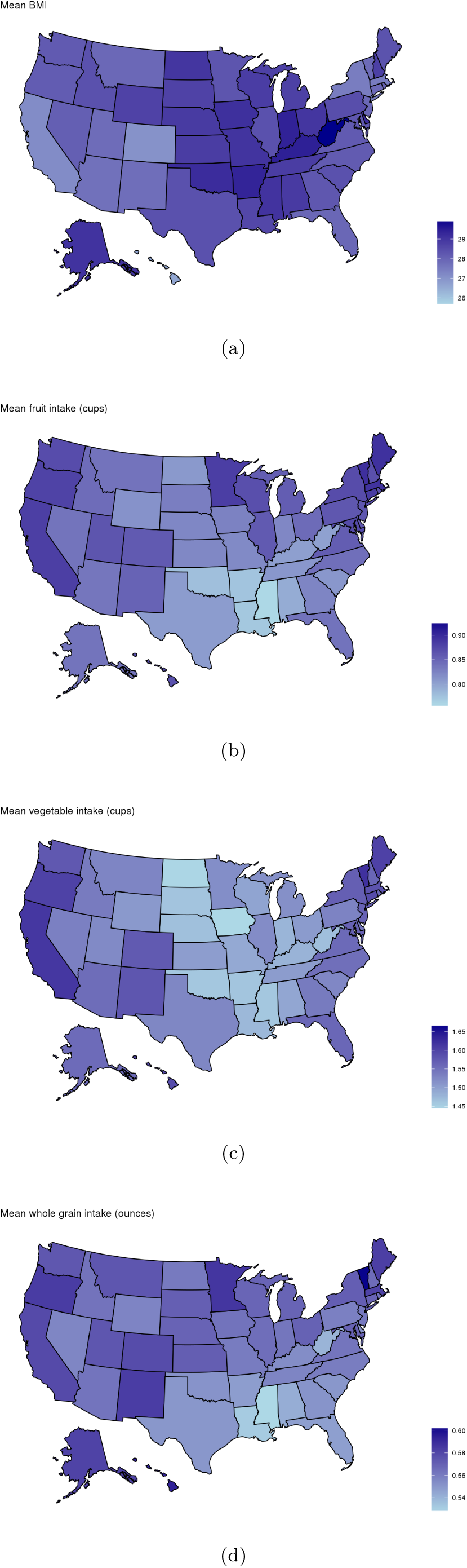
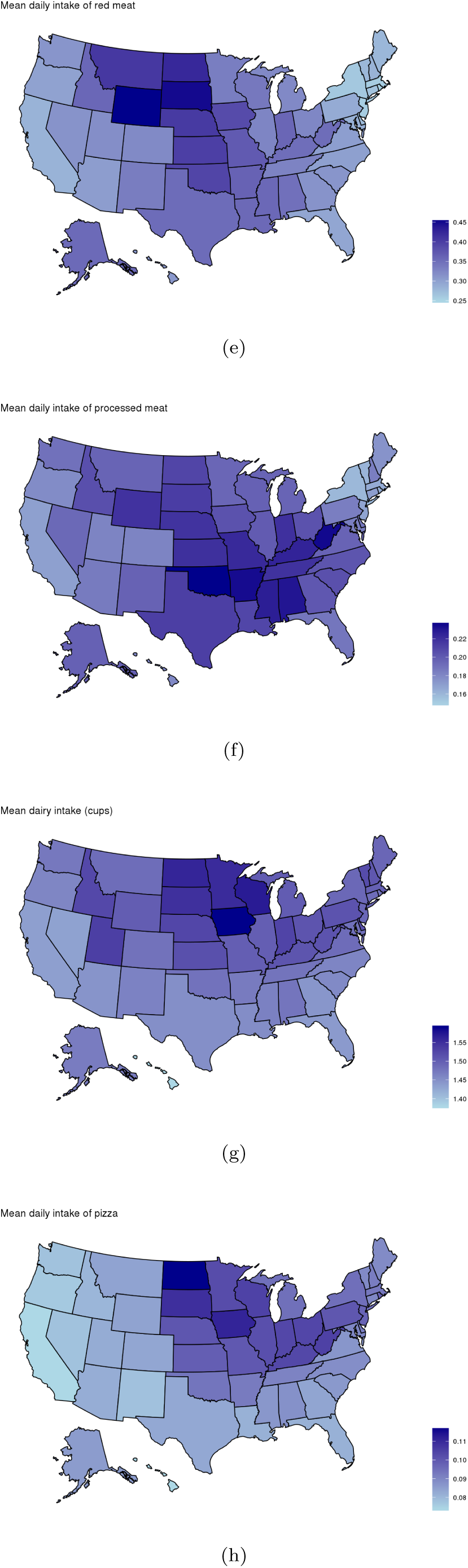
Maps of average body mass index, and average intake of select dietary factors by state, 2017-2019.

### Dietary intake estimates

Only 2.6% of the respondent population met national recommendations of fruit intake (1-½ -2 cups per day for adults depending on age and sex), with an average population intake of 0.83 cups/day.^(20)^ Mean vegetable intake (excluding French fries but including other potatoes) across all respondents was 1.54 cups per day, translating to only 5.9% of the total adult population who met guidance of 2-½ -3 cups/day depending on age and sex (Table 2 & 3). Vegetable intake showed a clear dose-response relationship with age, whereby among males and females aged 51 and over, 6.1% and 10.8% met recommendations, respectively. Among 20-30 year olds, only 1.1% of males and 0.3% of females met recommendations (Table 3). ^(21)^

**Table 2:**
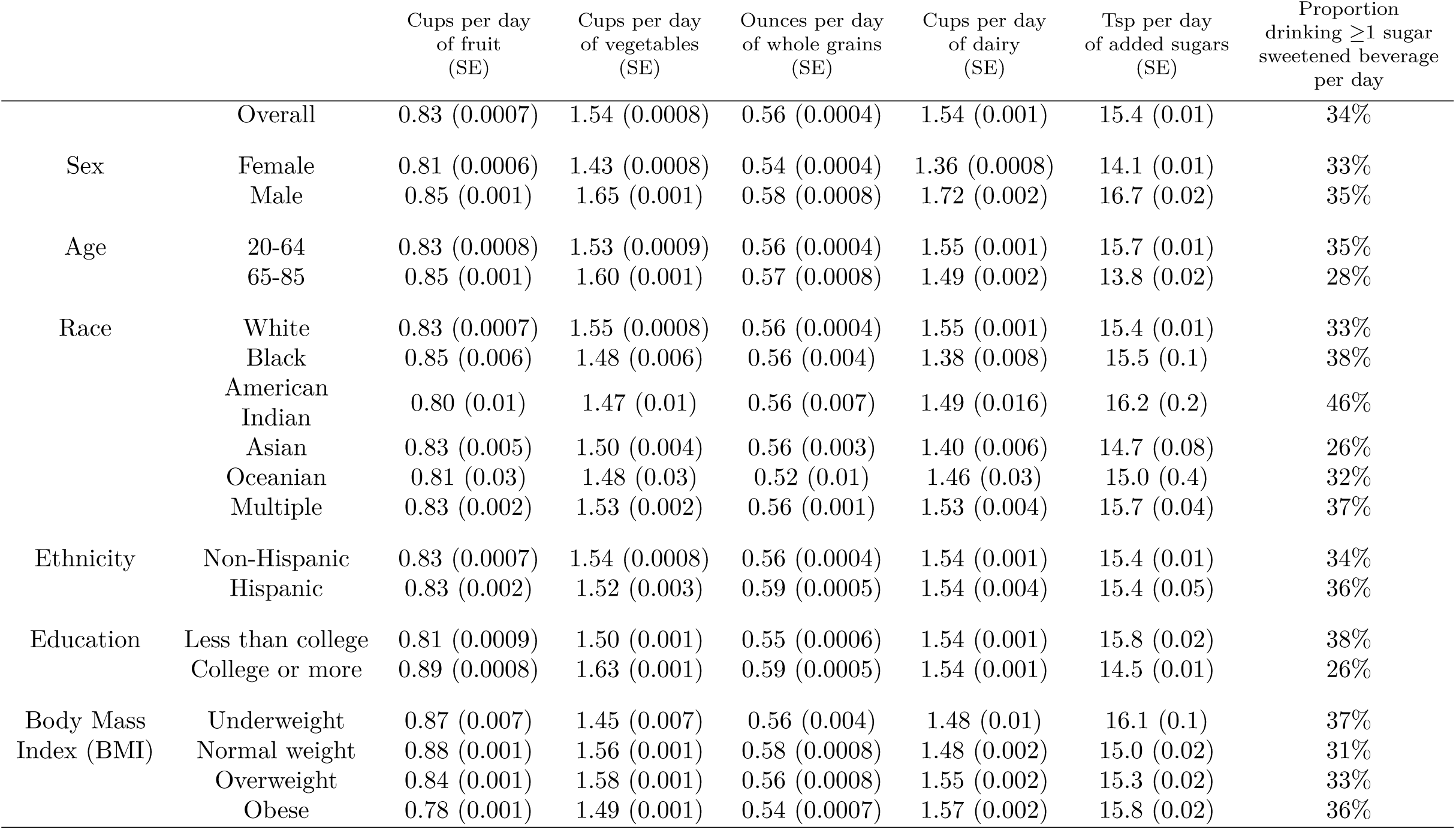
Respondent characteristics with complete data on age, sex, education, race/ethnicity, and body mass index compared to the national population drawn from the US Census.

| | | Cups per day<br>of fruit<br>(SE) | Cups per day<br>of vegetables<br>(SE) | Ounces per day<br>of whole grains<br>(SE) | Cups per day<br>of dairy<br>(SE) | Tsp per day<br>of added sugars<br>(SE) | Proportion<br>drinking $\geq 1$ sugar<br>sweetened beverage<br>per day |
| --- | --- | --- | --- | --- | --- | --- | --- |
|  | Overall | 0.83 (0.0007) | 1.54 (0.0008) | 0.56 (0.0004) | 1.54 (0.001) | 15.4 (0.01) | 34% |
| Sex | Female | 0.81 (0.0006) | 1.43 (0.0008) | 0.54 (0.0004) | 1.36 (0.0008) | 14.1 (0.01) | 33% |
|  | Male | 0.85 (0.001) | 1.65 (0.001) | 0.58 (0.0008) | 1.72 (0.002) | 16.7 (0.02) | 35% |
| Age | 20-64 | 0.83 (0.0008) | 1.53 (0.0009) | 0.56 (0.0004) | 1.55 (0.001) | 15.7 (0.01) | 35% |
|  | 65-85 | 0.85 (0.001) | 1.60 (0.001) | 0.57 (0.0008) | 1.49 (0.002) | 13.8 (0.02) | 28% |
| Race | White | 0.83 (0.0007) | 1.55 (0.0008) | 0.56 (0.0004) | 1.55 (0.001) | 15.4 (0.01) | 33% |
|  | Black | 0.85 (0.006) | 1.48 (0.006) | 0.56 (0.004) | 1.38 (0.008) | 15.5 (0.1) | 38% |
|  | American<br>Indian | 0.80 (0.01) | 1.47 (0.01) | 0.56 (0.007) | 1.49 (0.016) | 16.2 (0.2) | 46% |
|  | Asian | 0.83 (0.005) | 1.50 (0.004) | 0.56 (0.003) | 1.40 (0.006) | 14.7 (0.08) | 26% |
|  | Oceanian | 0.81 (0.03) | 1.48 (0.03) | 0.52 (0.01) | 1.46 (0.03) | 15.0 (0.4) | 32% |
|  | Multiple | 0.83 (0.002) | 1.53 (0.002) | 0.56 (0.001) | 1.53 (0.004) | 15.7 (0.04) | 37% |
| Ethnicity | Non-Hispanic | 0.83 (0.0007) | 1.54 (0.0008) | 0.56 (0.0004) | 1.54 (0.001) | 15.4 (0.01) | 34% |
|  | Hispanic | 0.83 (0.002) | 1.52 (0.003) | 0.59 (0.0005) | 1.54 (0.004) | 15.4 (0.05) | 36% |
| Education | Less than college | 0.81 (0.0009) | 1.50 (0.001) | 0.55 (0.0006) | 1.54 (0.001) | 15.8 (0.02) | 38% |
|  | College or more | 0.89 (0.0008) | 1.63 (0.001) | 0.59 (0.0005) | 1.54 (0.001) | 14.5 (0.01) | 26% |
| Body Mass<br>Index (BMI) | Underweight | 0.87 (0.007) | 1.45 (0.007) | 0.56 (0.004) | 1.48 (0.01) | 16.1 (0.1) | 37% |
|  | Normal weight | 0.88 (0.001) | 1.56 (0.001) | 0.58 (0.0008) | 1.48 (0.002) | 15.0 (0.02) | 31% |
|  | Overweight | 0.84 (0.001) | 1.58 (0.001) | 0.56 (0.0008) | 1.55 (0.002) | 15.3 (0.02) | 33% |
|  | Obese | 0.78 (0.001) | 1.49 (0.001) | 0.54 (0.0007) | 1.57 (0.002) | 15.8 (0.02) | 36% |

**Table 3:** Percent of the survey weighted 23andMe research participant population meeting federal dietary recommendations for fruits, vegetables, and dairy by age and sex (2017-2019).

|  |  | Fruit | Vegetables | Dairy |
| --- | --- | --- | --- | --- |
| Overall |  | 2.6 | 5.9 | 2.8 |
| Male | 20-30 | 4.2 | 1.1 | 7.7 |
|  | 31-50 | 1.6 | 1.6 | 5.5 |
|  | 51+ | 2.3 | 6.1 | 3.4 |
| Female | 20-30 | 0.4 | 0.3 | 1.1 |
|  | 31-50 | 3.7 | 8.4 | 0.9 |
|  | 51+ | 3.0 | 10.8 | 0.6 |

In stratified estimates of average intake, college education vs. no college education conferred higher intake of fruits, vegetables, and whole grains, with substantially less sugar intake both in total added sugars and the percent consuming at least one sugar sweetened beverage per day.

Differences across race/ethnicity showed Oceanian populations (i.e. Americans with origins in the Pacific islands) eat substantially less whole grains compared to other groups, and nearly half of American Indian respondents reported at least one sugar sweetened beverage per day. Dairy consumption was lower among Asians compared to other groups.

As shown in Figure 5, consumption of many of the food items included in the NHANES dietary screener show strong associations with BMI. In models adjusted for age, sex, education, and race/ethnicity, foods associated with the greatest increase in BMI included processed and red meat, fried potatoes, and pastry. Higher consumption of dairy products (cheese and ice cream) were all associated with higher BMI. Conversely, each tertile increase in the past month consumption of fruits, vegetables, whole grains, beans, and cereals corresponded to lower BMI. Frequency of chocolate consumption showed no statistically significant association with BMI.

**Figure 5:**
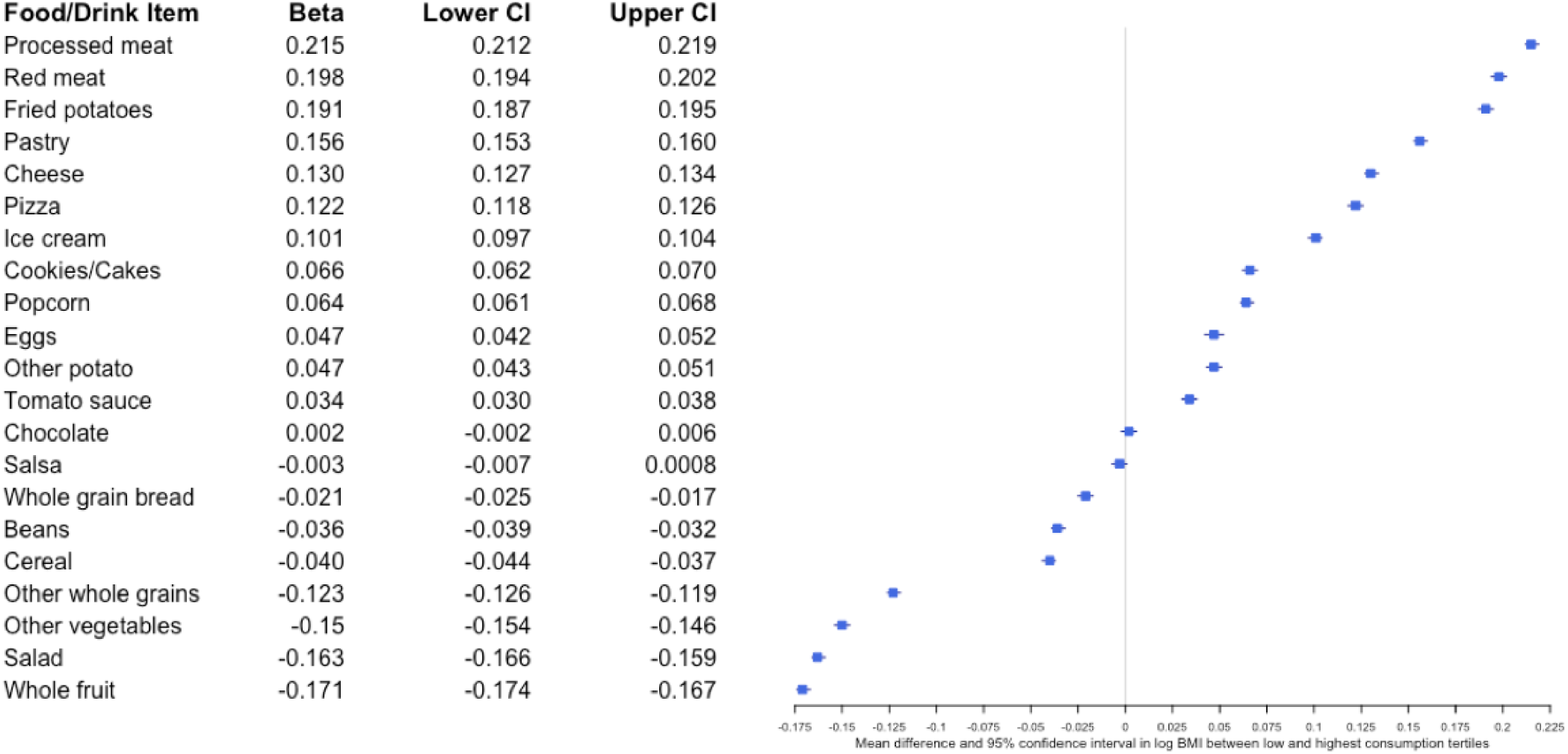
Mean difference in log body mass index (beta, 95% CI) between the highest and lowest consumption tertiles. Beta estimates and 95% confidence intervals were derived from linear models regressing intake tertile on log-transformed body mass index, adjusted for race/ethnicity, education, age (centered), sex, and age (centered)-squared.

## Discussion

Here, we detail demographic, temporal, and spatial characteristics of dietary factors measured by the DSQ and how they relate to BMI among 23andMe research participants. Because 23andMe research participants are twice as likely to have a college education, more likely to be female, white, and older than the general US population, dietary habits differential across these characteristics were most changed by weighting the sample to better represent the broader free living US population of adults. In the weighted sample, the majority of 23andMe participants fall well below dietary guidelines for consumption of fruits and vegetables showing remarkable similarity to the broader US population. In our stratified analyses, the most pronounced difference by education, which we used as a proxy for socio-economic status, was for soda consumption, which is less common as educational attainment increases. In our cross-sectional analyses of BMI and dietary habits, these under-consumed foods corresponded to lower BMI, whereas dairy, meats, and added sugars were associated with higher BMI. Descriptive spatial patterns of models of high vs. low intake of select dietary factors and BMI replicate results from large-scale nationally representative studies such as the Behavioral Risk Factor Surveillance System.^(23,24)^ Although it is generally agreed that there exists a troubling degree of bias in self-reported dietary intake data,^(25)^ observation of these patterns which are replicated across more rigorous data collection methods^(26)^ are reassuring.

Direct comparisons between weighted intake estimates among 23andMe research participants and the NHANES sample may be limited based on methodological differences in data collection. As described, 23andMe estimates are based on a brief screener, whereas the NHANES estimates are based on two in-person 24-hour food intake interviews.^(22)^ Comparing the survey-weighted results to the 2011-12 NHANES study, 23andMe respondents were most similar to the general U.S. population for fruits (0.73 vs. 0.69 median cups/day) and (1.54 vs. 1.63 cups/day) vegetables but less so for added sugars (15.4 vs. 18.5 tsp/day) and whole grains (0.56 vs. 1.00 ounces/day). Because of the significant time gap (5-6 year) between the most recently published NHANES values and the 23andMe data, it is possible that some amount of this difference is explained by the previously reported downward trends in consumption of added sugars observed over time.^(4)^

Measurement of dietary habits in the context of a self-reported web-based survey has the benefit of efficiency, but is hindered by the limited scope of data collection and its accuracy in reflecting the individuals’ general habits. Because web-based survey completion tends to decline as survey length increases,^(27)^ we chose the DSQ rather than a longer assessment such as a full food frequency questionnaire to maximize utility of the data collected with minimal respondent drop-off. However, the NHANES dietary screener (2009/2010)^(9,10)^ does not allow for estimation of total energy intake and may be a suboptimal tool for measuring the proportion of population meeting national fruit intake guidelines.

For example, due to a truncated response option at 2 or more times per day, for many adults (all men and women aged 19-30), it is not possible to report eating enough whole fruit to meet current dietary recommendations (2 cups per day for women aged 19-30, and men aged 19-50) if the portion size estimates are applied, which are less than 1 cup per serving. This becomes a greater problem for women, because the quantity multiplier (i.e. the estimated number of cups per serving) used to estimate cups per day is smaller for women than it is for men. Because the total fruit estimate is based on the combination of whole fruit and 100% fruit juice, participants that report the maximum serving size frequency (2 or more times per day) must also drink 100% fruit juice regularly to meet the recommended minimum fruit intake requirements.

Because fruit juice is not universally considered a healthful dietary option due to its high added sugar and low fiber content,^(28)^ those who eat fruit in quantities equal to or exceeding 2-cups per day but abstain from drinking fruit juice will be systematically misclassified by the DSQ as not meeting fruit intake recommendations. In an assessment of this screener, it was recommended to pilot test in each population prior to use, and to take caution when deriving precise estimates, but neither ceiling effects on fruit intake nor this particular problem was noted.^(11)^ Because the 23andMe respondent population includes an over representation of college educated people who we show here consume less added sugars compared to those without a college degree, limitations inherent to this questionnaire may be more pronounced.

Generalizability of the survey-weighted 23andMe respondent sample to the U.S. population was achievable for age, sex, education, white vs. non-white, and BMI. However, limits to sample weighting still require a higher representation of non-white participants to develop a generalizable weighted sample. An alternative approach would be sub-sampling the database^(29)^ to achieve a more balanced distribution across race/ethnicity, which we would recommend for analyses which aim to draw direct comparisons to the general population from the 23andMe research participant database. In addition to considerations of generalizability, other limitations to our study include the cross-sectional nature of the ascertainment for both exposure (diet) and outcome (BMI), which limits inferences of temporality and causality. Future studies may use prospectively collected data or Mendelian randomization analyses to further understand the causal architecture of our observed associations. Finally, our results are based on a one-time dietary measurement, which may not reflect habitual dietary intake.

In addition to the very low proportion of participants who met national guidelines for intake of fruits and vegetables, plots of reported intake over time are not encouraging. While some patterns are easily explained by broader trends, for example with milk, declines in reported consumption of fruits and vegetables over the study-period are either indicative of changing characteristics of the 23andMe research population or are reflective of broader declines in consumption of these foods.

Our exploration of nutrition patterns within the 23andMe database identified several unique advantages of this large-scale, participant-driven, digital cohort. The ability to quickly and contemporaneously collect nutritional information can inform more expeditious assessment of nutrition trends compared to traditional surveys; in contrast to the 23andMe data, NHANES data are usually made available to researchers several years after collection. Additionally, our unprecedentedly large sample size enables well-powered subgroup analyses, including future nutrigenomic studies, and rigorous ascertainment of spatial and seasonal variation in dietary intake. Finally, digital ascertainment of nutritional exposures minimizes participant burden, encouraging participation in future surveys.^(30)^

In conclusion, we have characterized the demographic, seasonal and spatial patterns of nutritional habits among 23andMe research participants in the U.S. Additionally, we report cross-sectional positive associations between BMI and the intake of red and processed meat as well as dairy, and inverse associations between BMI and the intake of fruit, vegetables, and whole grains. Our dataset offers a unique opportunity for rapid-scale real-time data collection, which can inform national trends in a much shorter time frame than current nationwide surveys. Efforts to diversify the 23andMe research participant database will increase the generalizability to the U.S. population, but because of the significant number of participants, survey sampling and weighting methods can achieve this at present. While large-scale cohorts like the 23andMe participants offer exciting future opportunities in precision nutrition, general efforts to continuously work towards improvements in dietary habits remain critical for maximizing health, maintaining a healthy BMI, and preventing chronic diseases.

## Data Availability

Data referred to in the manuscript are not available to the public.

## Figure Legends

Figure S1. Average intake reported by week of data collection, components of total dairy over two-year data collection period (2017-2019).

## Acknowledgements

We would like to thank the research participants and employees of 23andMe for making this work possible. Members of the 23andMe Research Team are: Michelle Agee, Adam Auton, Robert K. Bell, Katarzyna Bryc, Sarah K. Clark, Sarah L. Elson, Kipper Fletez-Brant, Pierre Fontanillas, Nicholas A. Furlotte, Pooja M. Gandhi, Karl Heilbron, Barry Hicks, David A. Hinds, Karen E. Huber, Ethan M. Jewett, Yunxuan Jiang, Aaron Kleinman, Keng-Han Lin, Nadia K. Litterman, Jennifer C. McCreight, Matthew H. McIntyre, Kimberly F. McManus, Joanna L. Mountain, Sahar V. Mozaffari, Priyanka Nandakumar, Elizabeth S. Noblin, Carrie A.M. Northover, Jared O’Connell, Steven J. Pitts, G. David Poznik, J. Fah Sathirapongsasuti, Anjali J. Shastri, Suyash Shringarpure, Chao Tian, Joyce Y. Tung, Robert J. Tunney, Vladimir Vacic, and Xin Wang

## Financial Support

This study was conducted with no external financial support.

## Conflict of Interest

All authors are employees of 23andMe, Inc., and hold stock or stock options in 23andMe. All authors had full access to all of the data in this study and take complete responsibility for the integrity of the data and the accuracy of the data analysis.

## Authorship

JS, RG, and SA formulated the research question. BC carried out statistical analyses. JS and SA wrote the article.

